# One year of testosterone therapy in a transmasculine amateur triathlete: changes in hormone cycles and exercise capacity

**DOI:** 10.1101/2025.09.11.25335594

**Authors:** Skyler R. St. Pierre, Sarah C. Johnson, Julie Muccini, Brianna Bourne, Benjamin H. Laniakea, Scott Delp, Jennifer Hicks

## Abstract

**Introduction:** The impact of testosterone therapy on hormone cycles, exercise capacity, and physiology of transmasculine individuals is not well understood. Existing studies report limited metrics at large time intervals between data collection.

**Methods:** Here, we collect high-resolution temporal data from a single amateur trans male triathlete over 13 months–including one baseline month and twelve months on testosterone therapy–to characterize hormone, strength, body composition, aerobic, and training load profiles.

**Results:** Daily urine hormone monitoring revealed that progesterone and luteinizing hormone are the clearest metrics associated with cessation of menses. After one year, the participant increased lean body mass by 12%, average hand grip strength by 16%, jump height by 16%, and average knee isometric strength by 18%, but did not lose fat mass or show changes in isokinetic knee strength. Fluctuations in running fitness and VO_2_ max were associated with both training and testosterone level with future work needed to untangle their relative contributions.

**Conclusion:** We provide guidelines to monitor trans males during testosterone therapy and recommendations to scale this case study to a larger population.

## Introduction

Sports matter. Participation in sports is associated with higher self-esteem and life satisfaction, and lower levels of depression, anxiety, and stress (1). Yet, trans people participate in sports at lower rates than cis people due to both *internal* barriers such as dysphoria and anxiety and *external barriers* including bullying, threats of violence, and sports policies (2). For trans people who desire physical changes to alleviate their dysphoria, gender-affirming hormone treatment leads to improved quality of life and mental health, significantly reducing depression risk (3, 4). Further, increased body satisfaction is the best predictor for increased physical activity for trans people (5), pointing to the need to support trans people’s access to gender affirming care *in parallel* with support for sports participation.

There is sparse research on trans men in sports (6), especially on how gender affirming testosterone therapy interacts with sport participation and fitness. Evidence from limited research on military personnel points to the capability of trans men on testosterone therapy to eventually match cis men’s athletic performance in standard military benchmarks but not exceed it (7). Trans men in the U.S. Air Force required 1-2 years after starting testosterone to reach the same average number of push-ups as cis men and 2+ years to reach the same mean 1.5-mile run time (8).

In non-athlete-specific populations of trans men, testosterone has been shown to increase lean body mass, grip strength, isometric and isokinetic strength, and decrease fat mass within one year (9, 10). Menstruation typically stops within 6 months of testosterone therapy, but if there is persistent menstruation, the reasons are often unclear (11). While there is speculation that testosterone dose and serum levels impact menstrual cessation, studies reporting high resolution hormone profiles over time are missing (11). Overall, the impact of testosterone gender-affirming care on physical performance for either trans athletes or non-athletes is under-researched, with limited population sizes and short monitoring durations (9).

Our study addresses two gaps in the research on transmasculine people in sports: the lack of (1) physiological data with frequent sampling in the first year of hormone therapy and (2) data across multiple modalities of performance and physiology for the same person. We hypothesize that simultaneous and high-frequency hormone, strength, and aerobic capacity monitoring will provide novel insight into the timing and magnitude of physiological changes as a result of testosterone therapy. We use historical training data to explore how training load and testosterone independently relate to fitness and performance in this case study.

## Materials and methods

### Study design

This study reports on a single transmasculine amateur triathlete. Active data collection occurred over 13 months as shown in **Figure 1**. The first month consisted of two baseline measurements, after which the participant started testosterone therapy. We also analyzed 1.5 years of the participant’s historical smartwatch exercise data. Dates from before the start of testosterone therapy are referred to as Pre-T, while dates during his first year on testosterone are demarcated with Year 1. The Stanford University ethics committee (IRB Protocol #72127) approved this study, and the study participant provided written informed consent for their voluntary participation in the research project and publication of the case.

**Figure 1.**
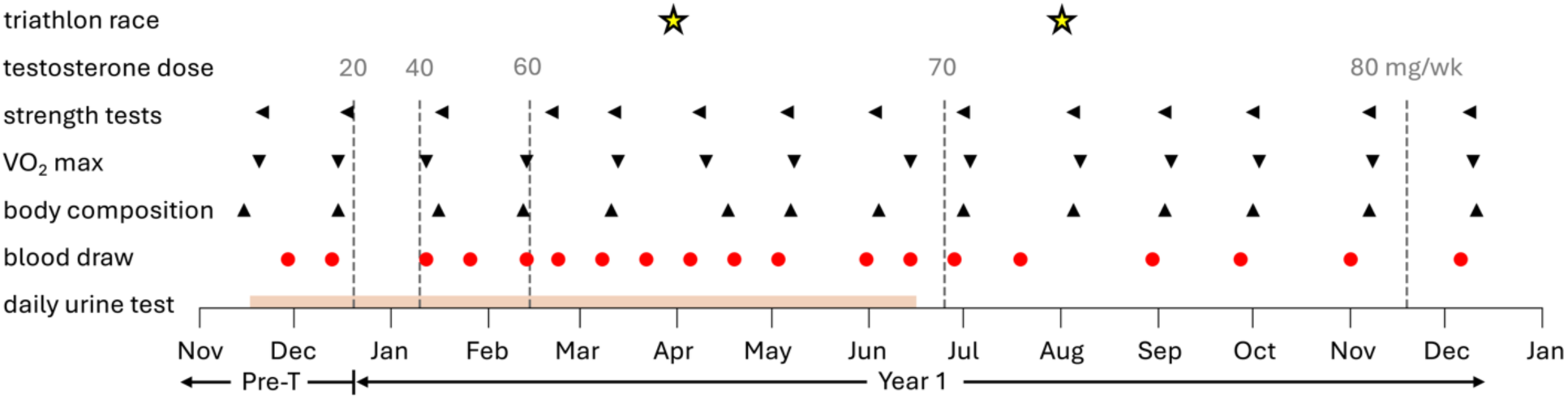
Timeline for active data collection. Active data collection included one baseline month and twelve months on testosterone therapy. The seven-month period of daily urine tests is highlighted in orange. Blood draws are shown as red dots and occurred twice a month for the first seven months and then monthly for the remaining time. Body composition, VO_2_ max, and strength tests were conducted monthly. Testosterone dosage level changes are marked by a dashed gray line with the total injection amount listed above in mg/wk. Stars show the two priority triathlon races during this period.

### Participant

The participant uses both he and they pronouns and will be referred to with both throughout this study. The participant is white and in his mid to late 20s. Further demographic details are withheld to protect the participant’s identity. He competed in three priority triathlon races with training, tapering, and recovery planned around these races in April Pre-T, April Year 1, and August Year 1. He self-reported menses and injuries. As this study is *n* = 1, we compare the results to minimum detectable changes (MDC) reported from prior literature. For all physiological testing, familiarization tests at easy to moderate effort were carried out with the participant one week prior to the initial test.

### Blood draws

For the first seven months of the study, the participant had their blood drawn at their local Labcorp (Laboratory Corporation of America Holdings, Burlington, NC) location approximately every two weeks. During the baseline month, two blood draws were taken, each measuring testosterone, estrogen, progesterone, luteinizing hormone, and follicle-stimulating hormone. For the next six months, to minimize blood volume drawn, one of two blood draws per month was used to measure testosterone, estrogen, progesterone, luteinizing hormone, and follicle-stimulating hormone and a second draw was used to measure testosterone only. After the first seven months, the participant underwent one monthly blood draw to measure all five hormones. After baseline testing, all blood draws except for one were taken mid-cycle, three days after the testosterone injection. That one exception was taken at trough, seven days after the injection. Progesterone values listed as <10, below the threshold of detection, were reported as 0 for plotting purposes.

### Testosterone therapy

The participant started testosterone therapy in December, by injecting testosterone cypionate subcutaneously at a dosage of 20 mg/wk. Based on bloodwork and clinical consultations, testosterone dose was increased by 10-20 mg/wk to achieve measured testosterone levels in the middle of the healthy cis male reference range of 300-1100 ng/dL (12).

### Urine sampling

We gave the participant an Inito (Inito, San Francisco, CA) ovulation kit to measure his estrogen, progesterone, luteinizing hormone (LH), and follicle-stimulating hormone (FSH) levels daily for the first seven months of the active data collection period. This smartphone collected reader has been validated against laboratory measurements (13). He collected the sample upon waking up except in a small minority of cases (<1%). We also instructed the participant to not overly hydrate but to maintain healthy hydration levels as best as possible. He missed three days in the seven-month period. For the analysis, we applied a Savitzky–Golay filter with a window length of 28, equivalent to one month, and polynomial order of 3 to smooth the raw data.

### Healthy cis female and male hormone reference ranges

We evaluated when the participant’s hormone levels shifted from average cis female to male hormone profiles. Healthy premenopausal cis female hormone levels are 15-46 ng/dL for testosterone (14) and 30-400 pg/mL for estrogen (15). The healthy cis male range for testosterone is 300-1100 ng/dL (12). The healthy cis male range for estrogen is reported as 6-50 pg/mL; however, estradiol levels are typically reported as a ratio with testosterone and few studies have looked at absolute estradiol levels in healthy cis men to inform this range (16). The healthy cis male reference ranges for LH and FSH are not well defined but generally span 1.8-13.4 mIU/mL for LH and 1.4-14.22 mIU/mL for FSH (16). Therefore, we establish a healthy male hormone profile as testosterone between 300-1100 ng/dL, estrogen between 6-50 pg/mL, LH between 1.8-13.4 mIU/mL, and FSH between 1.4-14.22 mIU/mL.

### Body composition

We measured the participant’s total body weight, body fat, and estimated resting metabolic rate via Bod Pod (COSMED, Concord, CA) every month. The Bod Pod is FDA-approved and uses Air Displacement Plethysmography to determine body composition (ratio of fatty mass to lean mass). The Bod Pod estimated resting metabolic rate using the Nelson equation (17). We set the sex of the participant to male for all measurements. Total energy expenditure was calculated as 2.08 times the resting metabolic rate using the built-in Bod Pod activity level factor for “very active,” corresponding to daily intense exercise. We instructed the participant to fast prior to the test and to only consume a moderate amount of water so as to be neither dehydrated or overhydrated. All tests were conducted in the morning. Although the Bod Pod is not yet validated in trans populations, consistent use of the same equation should capture relative changes from the baseline measurements.

### Leg and shoulder strength

We measured the participant’s joint torque strength (Nm) at the knee and shoulder joints using a Humac Norm isokinetic dynamometer (Computer Sports Medicine Inc., Stoughton, MA). The first test was a seated isometric hold of the knee extensors at 60 degrees of knee flexion. During each visit, the participant completed one practice test and then two repetitions at maximum effort. The second test was a concentric isokinetic movement for both knee flexors and extensors at a speed of 60 degrees/s. Again, there was a practice test, followed by five repetitions. The third test was the same as the second but at a rate of 180 degrees/s with 15 repetitions. Finally, the participant performed a standing internal/external shoulder rotation concentric isokinetic test (with elbow at 90 degrees of flexion) at a speed of 60 degrees/s for three repetitions after one practice test. All tests were repeated with both the left and right limbs, and we report the best repetition for each. An occupational or physical therapist conducted all of the testing. In November Year 1, the participant experienced a left deltoid strain (unrelated to the study) that affected his left shoulder strength and mobility for the last two time points.

### Countermovement jump height

To measure explosive power indirectly from leg strength, the participant performed three consecutive countermovement jumps with arm swing, recorded using OpenCap, a smartphone-based software to analyze human movement dynamics (18). We computed jump height as the difference between the OpenCap-determined maximum pelvis height and the average pelvis height while standing still over a several-second interval. We calculated mean and standard deviation across the three jumps. We calculated normalized jump height by dividing the mean jump height by the participant’s Bod Pod weight at each monthly time point. The participant performed no resistance training at any point in this study.

### Handgrip strength

To measure grip strength, the participant performed three repetitions of maximum squeezes on the Jamar hydraulic hand dynamometer for each hand with a small break between each test. He kept his elbow at 90 degrees and wrist in the neutral position while holding the dynamometer without resting it on his thigh and was instructed to keep his upper and lower body still and focus only on applying pressure to the dynamometer. We averaged the three efforts to compute the mean and standard deviation.

### Aerobic capacity: VO_2_ max

The participant underwent a monthly VO_2_ max test to analyze his energetic cost by way of metabolic gas exchange. His expired gases were measured breath-by-breath via a COSMED Quark Cardio Pulmonary Exercise Testing (CPET) (COSMED, Concord, CA) metabolic cart. The participant used an adjustable spin bike, which displayed watts for the test. The participant also provided an estimated Functional Threshold Power (FTP) from a test that used 95% of the maximum power output maintained for 20 minutes on a stationary bike. We used a standard step protocol for the V0_2_ max test consisting of a 10 minute easy spin followed by 4 minutes at FTP, a 3 minute break, then a 10 minute ramp testing starting from approximately 50% of FTP and increasing in 30 second intervals. In practice, this resulted in starting at 47% FTP, increasing 6% every 30 seconds up to a final power of 163% FTP. If the participant was able to hold the maximum power in the final ramp step, his FTP was increased by 15-20 watts for the following test to ensure he was hitting his maximum aerobic capacity within the ramp. These FTP increases happened in March and June. We recorded both *absolute* VO_2_ max (mL/min) and *relative* VO_2_ max (mL/min/kg). The participant’s weight was taken just prior to the test, and he was instructed to not eat three hours before the test. To analyze the data, we interpolated the raw results to evenly space the data to exactly every second and then used a moving window to find the best 30-second average VO_2_.

### Retrospective exercise data: Garmin smartwatch

The participant regularly recorded their training with a Garmin smartwatch device (Garmin Ltd., Olathe, KS) prior to the study start. We analyzed all recorded training data during the study as well as 1.5 years of prior data. We extracted total hours of exercise per month for running, biking, and swimming. Weekly indoor training rides ranging from 45-75 minutes were not recorded. We approximated the participant’s *fitness* by extracting the best 1500m running time each week normalized by his heart rate during that 1500m and reported the mean and standard deviation across four week intervals. We calculated the participant’s *performance* by extracting the best 800m, 1500m, 5 km, and 8 km running times each week and reported the mean and standard deviation across four week intervals. Elapsed time is used to ensure the participant was running for the entire duration rather than doing shorter intervals with breaks.

### Statistical analyses

We used Pearson’s linear correlations to explore the relationships between training stimuli, testosterone level, and aerobic capacity. We calculated weekly and monthly Training Impulse (TRIMP), adapted from the original Banister model which combined both intensity and duration of training, by using weighted activity intensity (using heart rate zones defined using ACSM guidelines (19)) × duration, summed across all sports (20). We correlated TRIMP and best recorded weekly running fitness (speed/HR) to serum testosterone and relative and absolute VO_2_ max and reported the Pearson correlation coefficient, *r*, and the p-value.

## Results

### Participant hormone levels shifted to cis male reference ranges within one year

Increasing testosterone dosage affected progesterone, estrogen, luteinizing hormone, and follicle-stimulating hormone levels, such that two months after starting testosterone therapy and at a dose of 60 mg/wk, all hormone concentrations remained within cis-male ranges through the conclusion of the study, **Figure 2**. At baseline, all hormones were within normal cis female ranges. The participant had testosterone levels of 23-60 ng/dL. No hormone data was collected at the first 20 mg/wk dosage of testosterone. At 40 mg/wk, the participant’s testosterone levels fluctuated between 265-347 ng/dL which is at the bottom or just below the healthy cis male range. At 32-34 pg/mL, estrogen remained within the healthy cis female range. Upon increasing his dosage to 60 mg/wk, testosterone rose to the middle of the healthy cis male range although testosterone levels continued to decline over the next four months towards the bottom of the cis male range. Just over one month after increasing dosage to 60 mg/wk, estrogen dropped below the healthy cis female range and stayed at 16-20 pg/mL, which is within the average reference ranges for cis men. Progesterone dropped below the Labcorp assay detection limit of 10 ng/dL at this dosage of 60 mg/wk. By mid-July, at 70 mg/wk, luteinizing hormone and follicle-stimulating hormone stopped fluctuating as noticeably and remained within 2.6-3.1 mIU/mL and 2-3.9 mIU/mL, within cis male ranges. All hormones were tested in serum once after increasing testosterone to 80 mg/wk with only testosterone level changing noticeably, rising to 513ng/dL.

**Figure 2.**
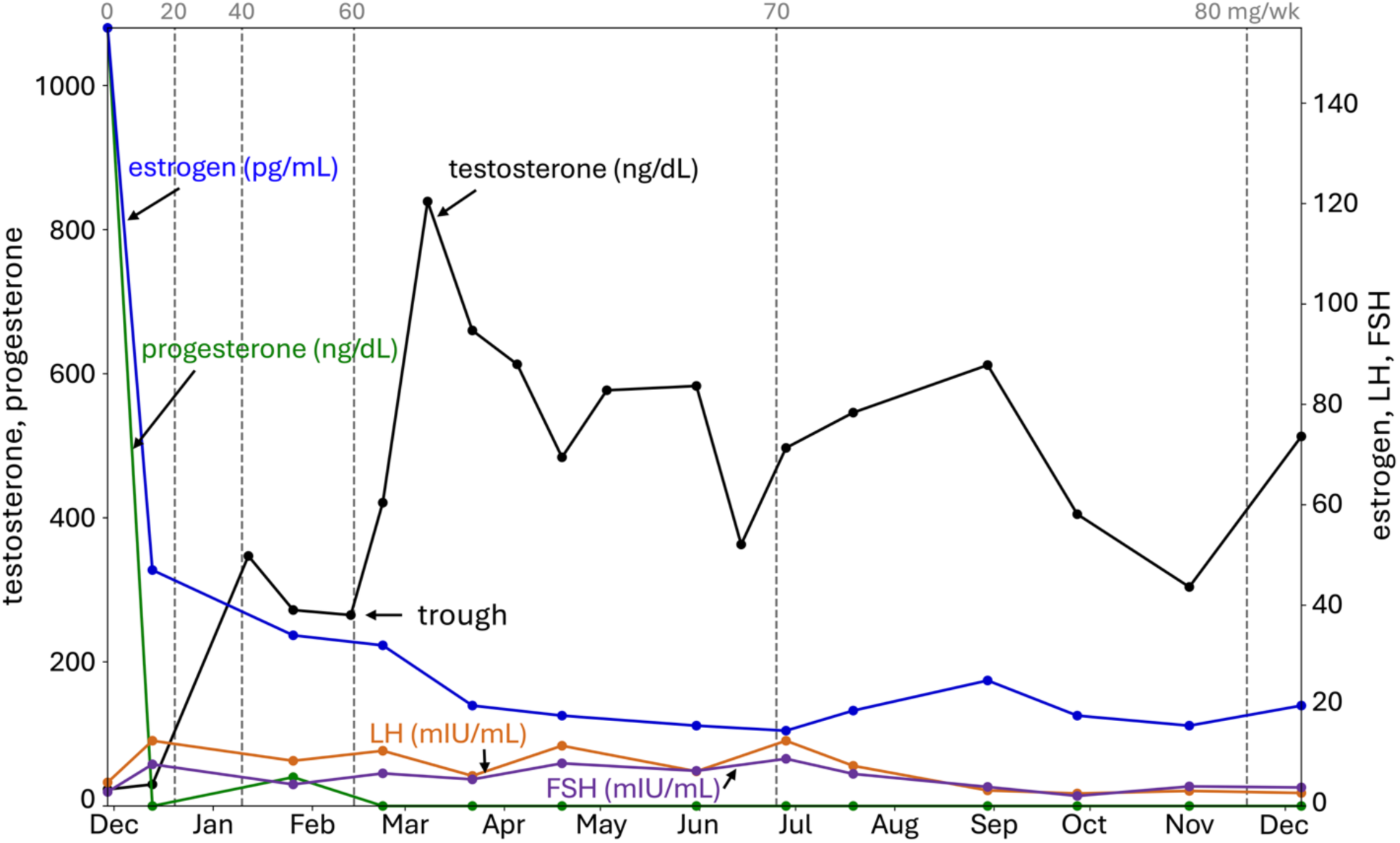
Testosterone, progesterone, estrogen, luteinizing hormone (LH), and follicle-stimulating hormone (FSH) measured via blood draws. Testosterone was measured every two weeks for the first seven months and then monthly for the duration of the study. In the baseline month, all levels were measured twice. Subsequently, all hormones except for testosterone were measured monthly. Testosterone levels were measured at midweek between injections (day 3) except for the time marked with trough on the graph where it was measured on day 7. Progesterone levels reported below the detection threshold of <10 ng/dL are shown as 0 on the graph. Testosterone dosage level changes are marked by a dashed gray line with the total injection amount listed above in mg/wk.

### Daily urine hormone monitoring reveals suppression of menstrual cycle associated with testosterone dose

The hormonal data collected via daily urine samples showed the impact of increasing testosterone dosage on gradual loss of characteristic hormonal fluctuations associated with ovulation and menses, **Figure 3**. During the baseline month and while at 20 mg/wk, the participant displayed peaks in luteinizing hormone (LH) and follicle-stimulating hormone (FSH) preceding peaks in E3G, the urine metabolite for estradiol, and PdG, the urine metabolite for progesterone that are indicative of normal ovulatory menstrual cycles. At 40 mg/wk, the raw data peaks in luteinizing hormone and E3G were both dispersed and suppressed, reaching 40% and 70% of their peak concentrations from the two menstrual cycles prior. Additionally, the temporal timing of estrogen rise following LH rise was much less pronounced and the increase in PdG that is due to secretion from the corpus luteum after egg release was no longer observed, suggesting ovulation had not occurred. Progesterone was the most impacted of the four hormones as PdG only reached 29% of the peak from the prior two menstrual cycles, while FSH was the least impacted as it reached 85% concentration. However, the participant still experienced menses this cycle. At 60 mg/wk, compared to 40 mg/wk, the smoothed data clearly demonstrated that the cyclic patterns of these four hormones were no longer coupled together. Although E3G continued to fluctuate, with a concentration range of 105-380 ng/mL, peaks in E3G were no longer in phase with peaks of LH or progesterone as they were pre-T. From one month after increasing dosage to 60 mg/wk to the end of the urine data collection in June, E3G, LH, PdG, and FSH were suppressed to 64%, 24%, 15%, and 71%, respectively, of their raw peak concentrations in the first two menstrual cycles. The last menses was just before the trough marker on **Figure 2**. The timeline for the cessation of menses was concurrent with all of the participant’s measured blood hormone concentrations staying within normal cis male ranges.

**Figure 3.**
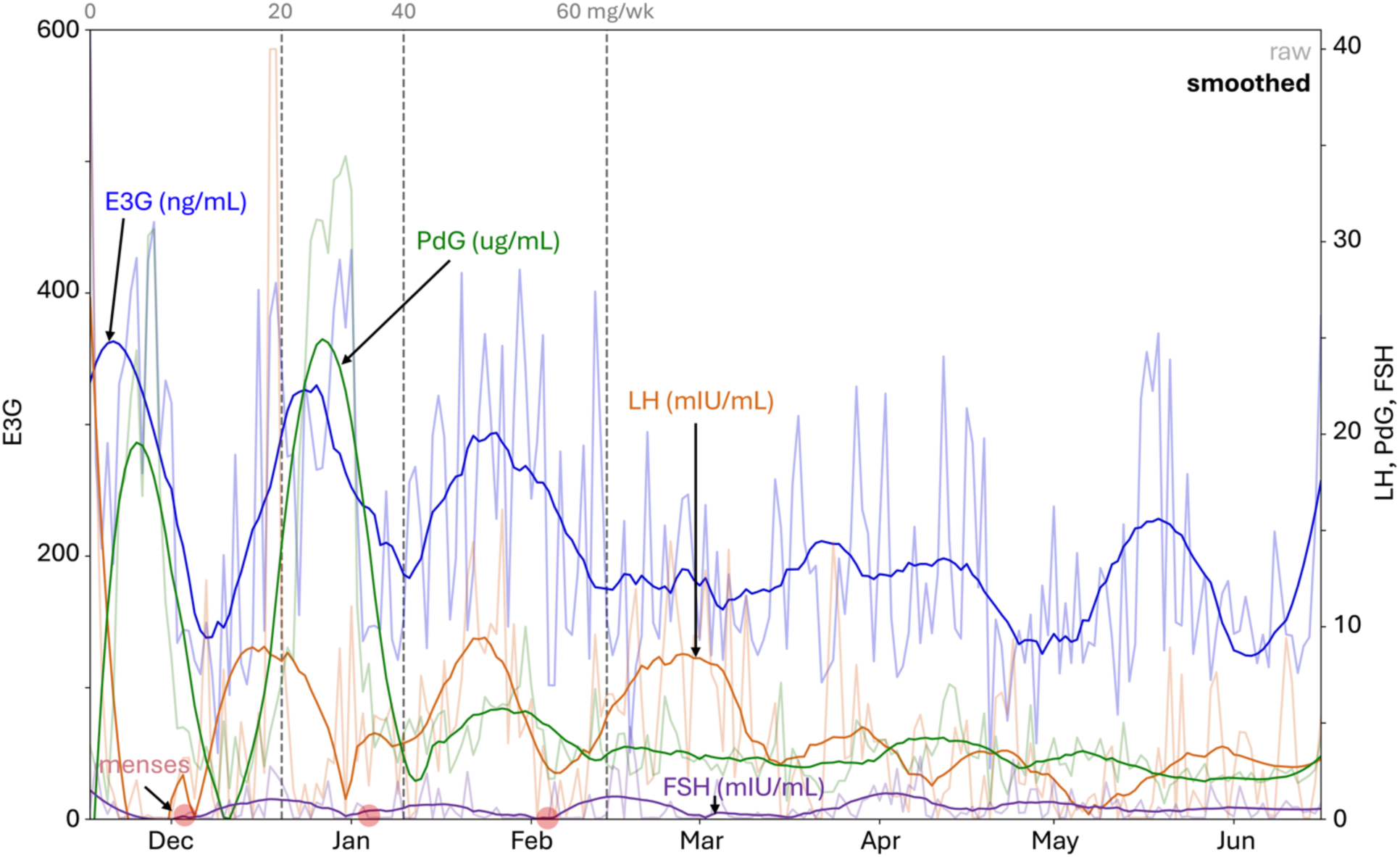
E3G, a metabolite of estradiol, luteinizing hormone, PdG, a metabolite of progesterone, and follicle-stimulating hormone measured daily for seven months using the Inito at-home urine testing kit. Raw data from the Inito app are shown in light shades and the smoothed data are shown in bold. Participant-reported menses are highlighted with a red dot on the x-axis. Testosterone dosage level changes are marked by a dashed gray line with the total injection amount listed above in mg/wk. Compared to the blood draw timeline in **Figure 2**, this urine data only encompasses the first seven months of the study.

### Testosterone treatment associated with linear increase in body weight

Upon starting testosterone, the participant’s total body mass increased 11% from 57 kg to 63.3 kg, with the fat free mass contributing 5.1 kg to the total increase and the fat mass contributing 1.2 kg, **Figure 4**, **Table 1**. Interestingly, the participant’s body fat percentage remained close to 25% for the duration of the study, yet he steadily gained weight at a rate of around 0.45 kg per month. The rate of fat free and total body mass increase was also remarkably consistent. The resting metabolic caloric needs of the participant increased by 137 kcal/day and their total energy caloric needs increased by 285 kcal/day.

**Figure 4.**
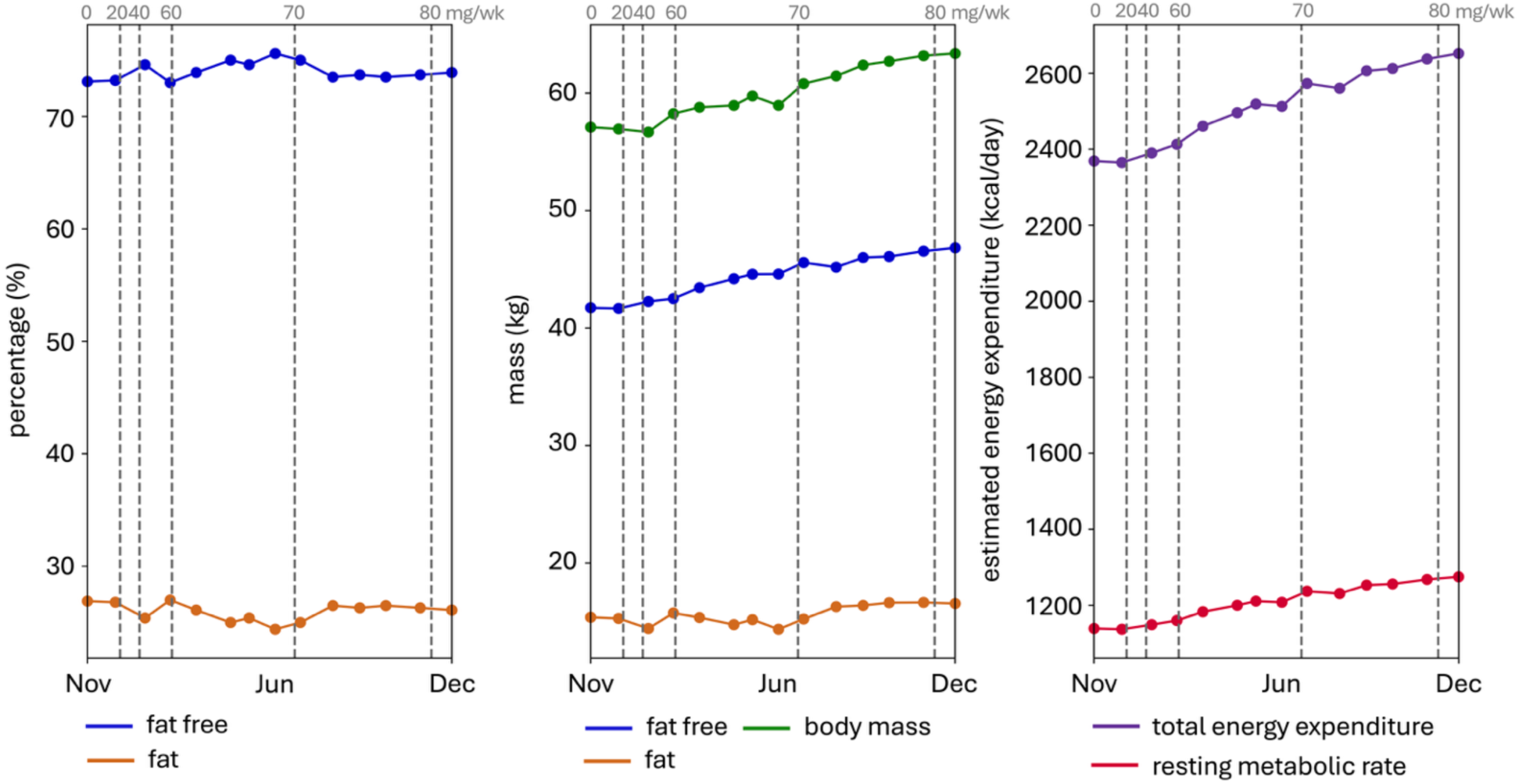
Body composition was measured via Bod Pod. The left figure shows fat and fat-free percentage. The middle figure separates fat mass, fat free mass, and total body mass, the sum of the fat and fat free mass. The right figure reports the estimated resting metabolic rate calculated by the Bod Pod as well as the total energy expenditure with a chosen activity level of “very active” corresponding to an intense workout every day. Testosterone dosage level changes are marked by a dashed gray line with the total injection amount listed above in mg/wk.

**Table 1.** Baseline and one year of transition serum hormone, body composition, aerobic capacity, jump height, and strength characteristics. We average the two baseline measurements, left and right sides, and multiple trials when relevant and report the mean ± standard deviation.

|  | Baseline | 1 Year |  | Baseline | 1 Year |
| --- | --- | --- | --- | --- | --- |
| Testosterone (ng/dL) | 27 ± 5 | 513 | Grip strength (kg) | 28.1 ± 2.0 | 32.7 ± 1.2 |
| Estrogen (pg/mL) | 101 ± 76 | 20 | Jump height (cm) | 29.4 ± 0.5 | 34.2 ± 1.0 |
| Progesterone (ng/dL) | 1080, <10 | <10 | Knee isokinetic extension 60 deg/s (Nm) | 82.8 ± 5.2 | 89.5 ± 14.8 |
| FSH (mIU/mL) | 5.6 ± 3.9 | 3.7 | Knee isokinetic flexion 60 deg/s (Nm) | 51 ± 4.3 | 53 ± 1.4 |
| LH (mIU/mL) | 8.9 ± 5.9 | 2.6 | Knee isokinetic extension 180 deg/s (Nm) | 59.8 ± 4.6 | 64.5 ± 7.8 |
| Fat free mass (kg) | 41.7 ± 0.0 | 46.8 | Knee isokinetic extension 180 deg/s (Nm) | 39.8 ± 3.2 | 44.0 ± 1.4 |
| Fat mass (kg) | 15.3 ± 0.1 | 16.5 | Knee isometric extension (Nm) | 111 ± 11 | 131 ± 14 |
| Absolute VO<sub>2</sub> (mL/min) | 2837 ± 48 | 3159 | Shoulder isokinetic internal rotation 60 deg/s (Nm) | 19 ± 1.6 | 22 ± 4.2 |
| Relative VO<sub>2</sub> (mL/min/kg) | 49.1 ± 1.4 | 49.6 | Shoulder isokinetic external rotation 60 deg/s (Nm) | 10.5 ± 2.1 | 13 ± 1.4 |

### Changes in lower and upper body strength were unclear after one year of treatment

We did not observe a clear trend in knee isokinetic strength (**Figure 5(a)** and **5(b))**. In contrast, the participant’s knee isometric extensor strength increased in both the right and left leg from the average baseline measurements by 21 Nm (17.5%) and 19 Nm (18.6%) at one year, respectively, although with very large fluctuations over the duration of the study, **Figure 5(c)**. We again do not see substantial changes in shoulder strength, but note an upward trend in right internal shoulder rotation strength, **Figure 5(d)**, **Table 1**.

**Figure 5.**
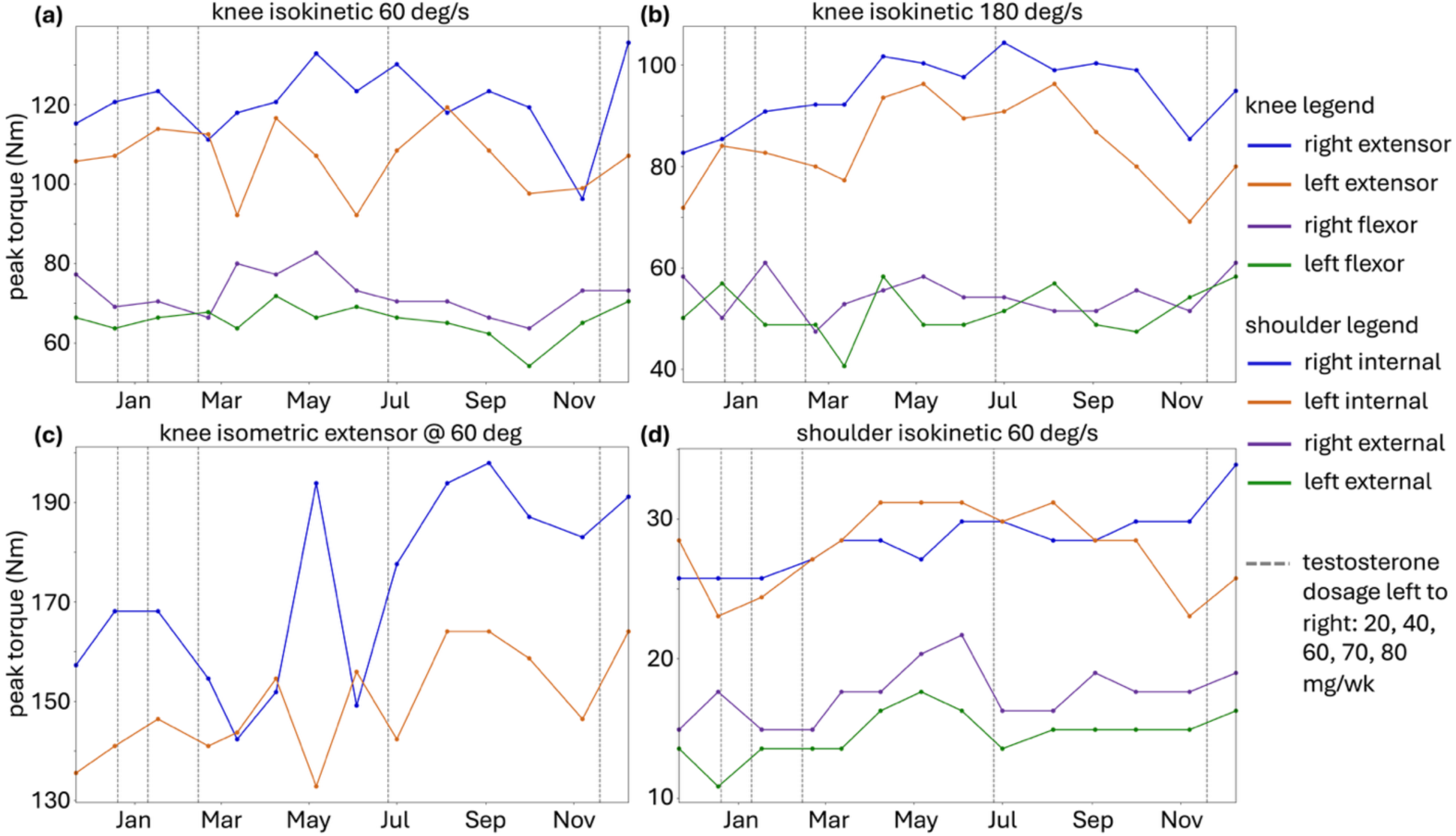
Peak torque about the knee and shoulder tested monthly using a Humac Norm isokinetic dynamometer. Tests were performed with isometric or concentric muscle contraction: (a) shows peak torque about the knee with isokinetic motion at 60 deg/s and (b) is the same test but at 180 deg/s; (c) shows peak torque for a knee isometric hold at 60 degrees; (d) shows peak shoulder torque at 60 deg/s. Testosterone dosage level changes are marked by a dashed gray line with the total injection amount listed above in mg/wk.

### Grip strength and jump height increased after one year of treatment

Absolute jump height increased 4.8 cm (16%) from baseline to Year 1 in December, **Figure 6**. The participant’s jump height normalized by body weight peaked in June and July and then decreased to close to the baseline as his weight increased. Although the participant did not train grip strength, we observed that his right and left hand grip strength increased, albeit non-monotonically, **Figure 7(a)**. At the final time point, the participant’s average right and left hand grip strength had increased by 4.6 kg or 16%.

**Figure 6.**
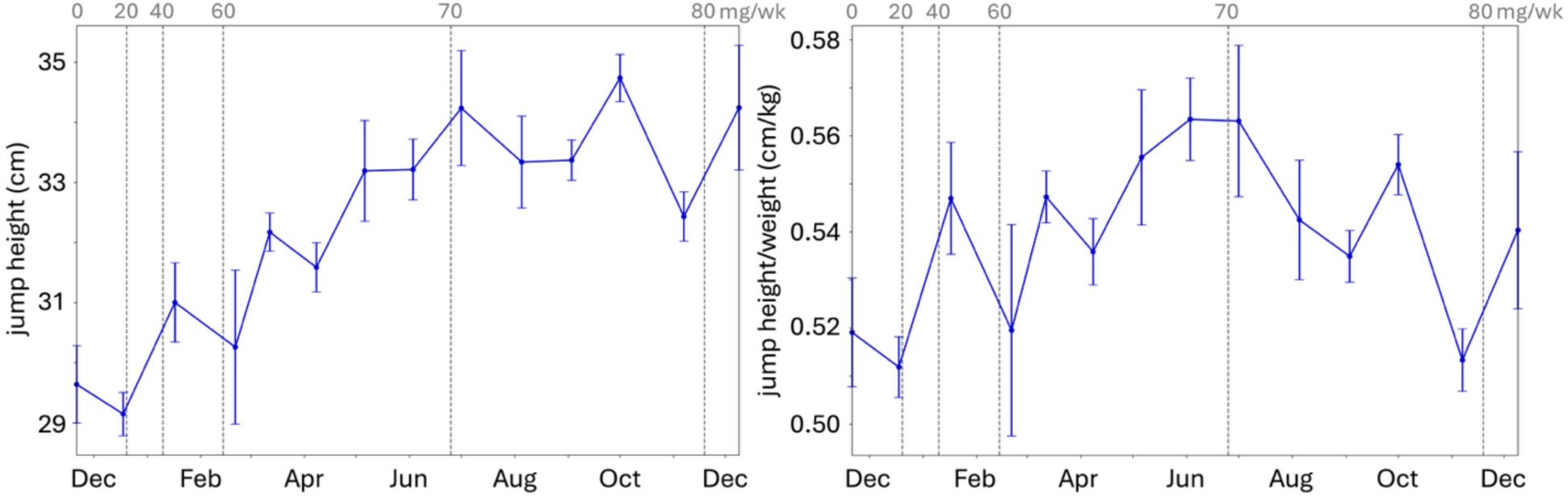
Maximum jump height and jump height normalized by body mass measured using a countermovement jump. The participant performed three countermovement jumps with a small rest in between jumps. The mean and standard deviation are shown. The left graph reports the jump height while the right graph reports jump height normalized by the participant’s body weight from the most recent Bod Pod test. Testosterone dosage level changes are marked by a dashed gray line with the total injection amount listed above in mg/wk.

**Figure 7.**
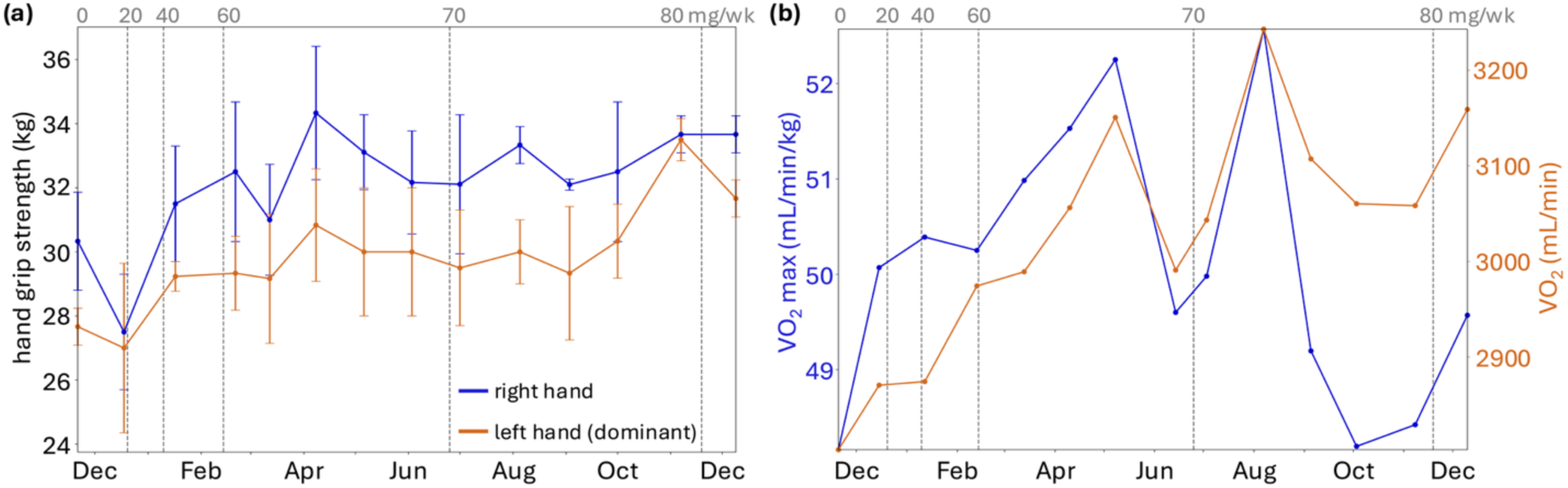
(a) Hand grip strength measured by Jamar dynamometer. The participant’s left hand is his dominant hand. He performed three grip tests on each hand monthly. The mean and standard deviation are shown. (b) Maximum and absolute oxygen consumption measured monthly using a 10-minute ramp test on a stationary bike with a metabolic analyzer. The left axis shows VO_2_ max normalized by body mass while absolute VO_2_ max is reported on the right axis. Testosterone dosage level changes are marked by a dashed gray line with the total injection amount listed above in mg/wk.

### Aerobic capacity, Training Impulse, and testosterone level showed significant correlations

The participant’s relative VO_2_ max (mL/min/kg) peaked twice during the study duration but ultimately did not increase from baseline by the end of the study, **Figure 7(b)**. Relative VO2 max (mL/min/kg) was not correlated with Training Impulse (TRIMP) as determined from the participant’s smartwatch data (p = 0.073, r = 0.49) but was significantly correlated with testosterone levels (p = 0.020, r = 0.63), **Figure 8**. Interestingly, absolute VO_2_ max (mL/min) did increase by 322 mL/min (11%) from baseline to the last time point at the end of Year 1, **Table 1**. Absolute VO2 max (mL/min) was not correlated with TRIMP (p = 0.140, r = 0.41) but was significantly correlated with testosterone (p = 0.006, r = 0.71). Running fitness extracted from the smartwatch data (speed/heart rate) was significantly correlated with TRIMP (p = 0.004, r = 0.27) but not correlated with testosterone levels (p = 0.122, r = 0.45), **Figure 8**.

**Figure 8.**
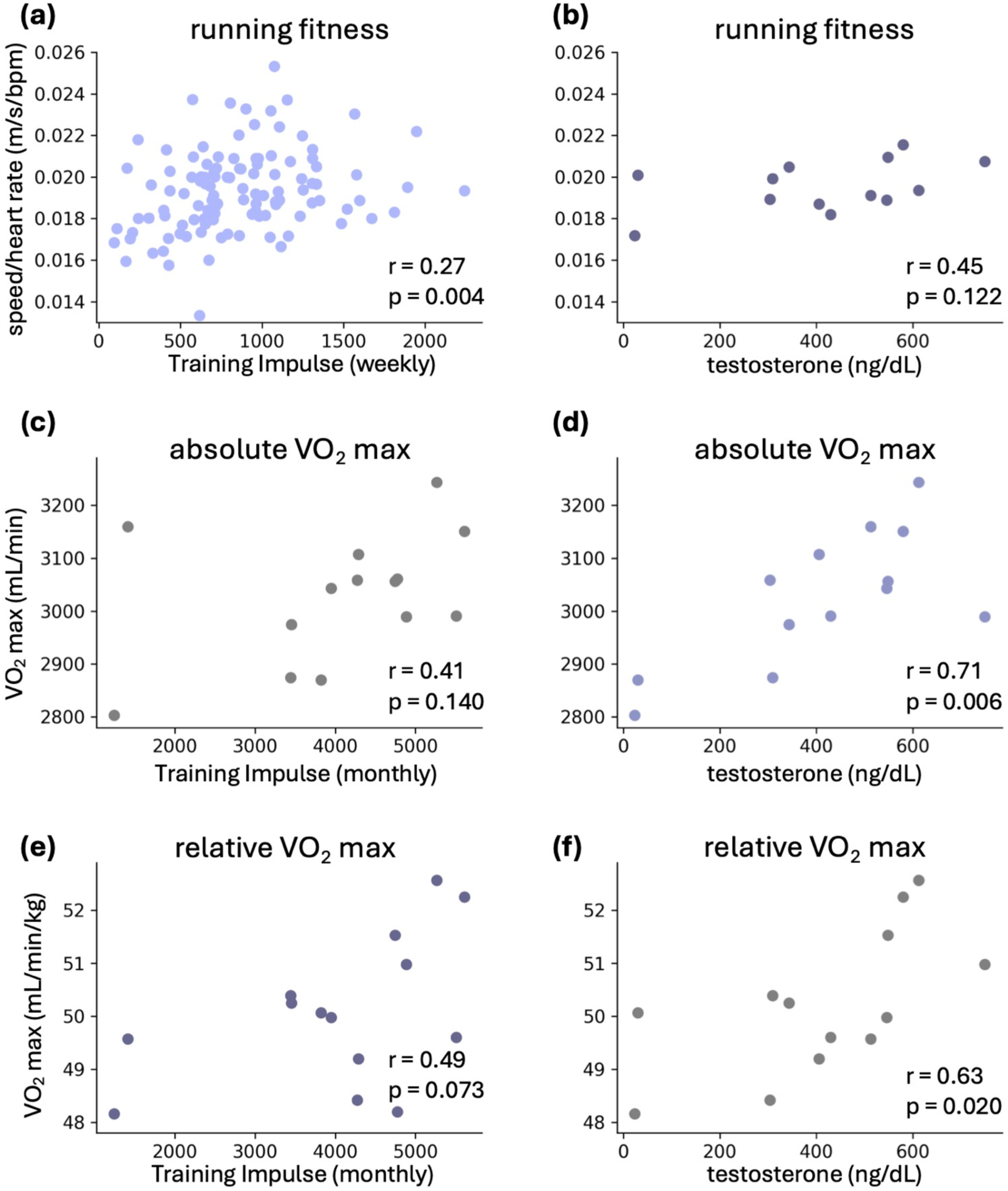
Correlations between running fitness, aerobic capacity, Training Impulse, and serum testosterone levels. Top row: speed/heart rate for fastest weekly 1500m versus (a) weekly Training Impulse (TRIMP) across swimming, biking, and running and (b) monthly testosterone levels. Middle row: Absolute stationary cycling V0_2_ max versus (c) monthly TRIMP and (d) monthly testosterone levels. Bottom row: Relative V0_2_ max (absolute V0_2_ max normalized by bodyweight) compared to (e) monthly TRIMP and (f) monthly testosterone levels. We report the Pearson’s correlation coefficient, *r*, and p-value for each plot.

The first peak in VO_2_ max in May did not directly correspond with either of the two priority races carried out during the study, but may reflect the fitness gained following the first race in the prior month, in addition to the start of increased exercise volume leading up to the second race in August. The second peak in VO_2_ max coincides with the timing of this second race. Running fitness extracted from the smartwatch showed two peaks that consistently aligned with annual April priority races both on and off testosterone, **Supplemental Figure 9**, **SDC**.

We also used the smartwatch data to extract fastest running speeds across various distances with minimal elevation change, as a direct marker of his *performance* over time, **Supplemental Figure 10**, **SDC.** We found that raw speed varied collectively across 800m, 1500m, 5km and 8km distances such that all speeds improved or decreased together. There appeared to be only marginal differences in speed between the year prior to and the year on testosterone. However, the participant increased both his run volume and consistency right around the time he started testosterone which confounds interpretation of the results.

## Discussion

This study presents high-resolution temporal data across the first year of testosterone therapy from a single transmasculine recreational athlete and provides novel insight into hormone cycles, exercise capacity, and muscular physiology.

### The effect of testosterone dosage on hormone cycles is evident with daily hormone measurements, but not with once or twice monthly tests

The current standard of care for transmasculine people taking testosterone therapy is to measure serum testosterone levels via blood draws every 3 months in the first year, and then once or twice yearly afterwards (21). As shown in **Figure 2**, for this participant such infrequent blood draws would delay the identification of his decline in testosterone levels below the healthy cis male range. More frequent blood draws may allow patients seeking rapid masculinization to adjust their dosages sooner to reach cis male hormone ranges.

Although testosterone concentration is known to play the largest role in bringing about masculinizing effects, trans men are also interested in knowing when their menses will stop as this can be a large contributor to dysphoria (22). High frequency urine testing was able to reveal *when* the addition of exogenous testosterone started to suppress and disrupt the participant’s original hormone cycles and *which dosage* led to a new sustained hormone profile without menstrual cycling, **Figure 3**. Notably, the urine metabolites for progesterone and luteinizing hormone provided a clearer indicator than estrogen for the end of cis female hormonal patterning which in this case was associated with cease of menses. Contrastingly, estrogen concentrations maintained large fluctuations between 105-380 ng/mL in the last month of the seven month monitoring period.

The participant ceased menstruating when all hormones, via both blood serum and urine testing, sat within cis male levels. This happened at a dosage of 60 mg/wk, two months after starting testosterone treatment. Most trans men stop menstruation within six months (23). Having greater knowledge of daily hormone fluctuations could provide valuable insight for trans men with persistent menstruation and inform clinicians if the solution is as simple as increasing testosterone dosage or if there are other factors (11, 24).

### Testosterone was associated with steady weight gain at a lower body fat ratio than baseline, but overall body fat percentage decreased only marginally within a one-year time period characterized by consistent exercise

The participant gained 11% or 6.3 kg of total body mass in one year, split into an increase of 5.1 kg of fat free mass and 1.2 kg increase in fat mass, **Figure 4**. The increase in fat free mass was larger than the minimum detectable change (MDC) of 1.87 kg (25). Interestingly, we found that weight gain was nearly constant every month, which no other study has examined. Previously reported increases in mass range from 2.5 – 5.3 kg (26–28). Overall, the data from our participant align with the general consensus that testosterone treatment will increase lean body mass and total body mass (29).

Several studies report substantial decreases in fat mass of around 2.5 kg over one year on testosterone (26, 30); however, the fat mass of our participant increased slightly, resulting in only a 0.8% decrease in body fat percentage over one year. Notably, our participant had top surgery before starting hormone therapy which may confound direct comparisons with prior studies as testosterone causes a significant reduction in breast tissue fat to levels comparable to postmenopausal cis women (31). There is very limited data on expected body composition changes for athletic trans men specifically. Roberts et al. found that after one year on testosterone the total body mass of trans men in the U.S. Air Force increased by 3.7% or 2.6 kg but after two and a half years their body mass was nearly back to baseline (8). Clearly, more research is needed to understand the body composition changes of athletic trans men, for longer time periods, and with larger populations. Importantly, the equations used to calculate the Bod Pod’s body composition and caloric metrics have not been validated in trans cohorts and in general have significant error at the individual level so should be cautiously interpreted (17). Varying hydration levels may also increase this error.

### Knee isometric strength, jump height, and hand grip strength all increased in the absence of training focused on these tasks

Several studies, albeit many with small sample sizes and most on non-athletic populations, have investigated the effect of testosterone on muscular strength. Increases in muscle mass and decreases in subcutaneous fat are expected as a result of taking testosterone therapy, leading to greater muscle strength (32). Our participant gained on average 20 Nm or 18% for knee isometric extension at 60 degrees, **Figure 5**. This change was smaller than the reported MDC of 51.5 Nm (33) but comparable to the changes observed by Wiik et al. While Wiik et al. also showed increases in strength for isokinetic strength in extension and flexion (10), our participant showed fluctuation over time rather than a clear trend. However, **Table 1** reveals that the average of right and left sides did increase at least marginally for all strength metrics. Further work examining the timeline of isokinetic versus isometric changes in trans men is needed.

One study of 12 trans male athletes who had been on testosterone therapy for at least one year found that the jump height of trans men was not statistically different from cisgender male athletes (34). Our participant demonstrated increases of 16% (4.8 cm) for jump height, **Figure 6**, higher than the MDC of 1.5 cm via motion capture (35). Future work with larger populations is needed to investigate the average effect of testosterone on jump height.

Grip strength is expected to increase in trans men (26, 36). Our participant had an average increase of 16% (4.6 kg) in hand grip strength, **Figure 7(a)**, but this was less than the MDC of 6 kg (37). As observed by Scharff et al., (2019), our participant had the biggest changes in grip strength in the initial months of testosterone therapy and then a noisy plateau afterwards.

### Increased body weight limits increases in relative VO_2_ max

Similarly to jump height, no studies have examined changes in VO_2_ max in trans men upon starting testosterone, although in cisgender women low dose testosterone cream was shown to increase the total time to running exhaustion compared to a placebo (38). Hamilton et al. found that athletic trans men who had been on hormone therapy for at least one year had lower absolute VO_2_ max (mL/min) than cis men, but comparable relative VO_2_ max (mL/min/kg) (34). As seen in **Figure 7(b)** and **Table 1**, our participant had an increase of 11% (322 mL/min) in absolute VO_2_ max, which was larger than the reported MDC for cycling of 100-170 mL/min (39), but did not improve from baseline by the end of the study for relative VO_2_ max. Because the participant gained 6.3 kg total after 12 months on testosterone, he would need to be proportionally more aerobically fit to hit the same relative VO_2_ max as at baseline with a lower weight, aligning with known scaling principles.

Our preliminary statistical analyses suggest that both testosterone and increased training improved aerobic capacity, **Figure 8**, but further work is needed to determine their independent contributions. A major limitation of this study is that our participant increased overall training load following testosterone treatment. Future work should also investigate the relative impact of cumulative time on testosterone versus serum testosterone levels on physiological and fitness changes.

### Recommendations for scalability: insights gained from our high temporal resolution, multi-factor, thirteen-month n=1 study

Our participant spent on average 8 hours per month completing data collection. For 6 of those hours, an occupational therapist oversaw and ran the various tests. Thus, scaling every component of this study to a larger population would require prohibitive participant and staff time. Based on participant feedback, our results, and the literature, we would recommend reducing Bod Pod, Humac Norm strength and VO_2_ aerobic testing to less frequent time intervals than monthly. While most body composition, strength and aerobic performance changes happen within the first 1-2 years on testosterone (9), the resources, effort, and time required to do monthly measurements outweighed the benefits of added information. We believe that a 3-6 month interval would have provided a good balance between adequate data and less participant and staff burden.

Grip strength and jump height are quick, informative measurements. Depending on the goal of a study, choosing intervals between monthly and every 2-3 months would still be informative without missing the plateau phase that can occur with grip strength within the first 6 months (36). Additionally, the open-source OpenCap software system requires only two iPhones and tripods, making recording jump height as well as full-body musculoskeletal kinematics in larger participant populations efficient and accessible (18).

Further research is needed to determine optimal hormone testing frequency for trans athletes. Based on our case study data, we recommend monthly blood draws, for those comfortable with needles, during the first 6 months. Daily urine monitoring may be particularly beneficial for studies examining persistent menstruation in trans men. However, more research is needed to examine whether high-frequency monitoring affects the transition timeline of trans men in larger populations. Additionally, advances in wearable technology to continuously measure sex hormones would be of significant benefit to both trans and cis people (40).

Finally, passive training monitoring via smartwatch data is the easiest metric to scale to large populations and should be explored further for trans men and trans athletes in order to both control for training load and understand the positive effects of training on desired strength, fitness and body composition outcomes. Diet and sleep monitoring may also provide novel insight into the physiological impacts of testosterone therapy.

## Conclusion

We provide the single-most comprehensive study of one amateur trans male triathlete for a total of 13 months across hormone, strength, body composition, aerobic, and training metrics. The daily hormone monitoring revealed the effects of testosterone dosage that the standard of care would traditionally overlook. In line with prior studies, our participant gained lean body mass, but unlike prior studies, did not lose fat mass. As expected, hand grip strength and knee isometric strength increased. For the first time, we also looked at jump height and VO_2_ max and found that while jump height increased steadily over time, fluctuations in VO_2_ max were associated with both Training Impulse and testosterone level. Our study provides recommendations to scale up to a larger study population and offers potential guidelines to monitor hormone therapy in the trans population.

## Positionality Statement

The first author, SS, is a trans man. He designed and conducted this study to answer questions around testosterone therapy that he saw as most pressing in his own community: namely, what changes will happen and when. BL is also trans and is a doctor practicing in LGBTQ+ health. They provided expert guidance in designing and interpreting this study to help inform the clinical practice of gender-affirming healthcare. All authors are committed to supporting research on, with, and by trans people (41, 42).

## Human Subjects Statement

The Stanford University ethics committee (IRB Protocol #72127) approved this study, and the study participant provided written informed consent for their voluntary participation in the research project and publication of this case.

## Acknowledgements

The authors thank Vien Vu for his help with the Humac Norm experiments and Kristen Gravani for her help with the Bod Pod experiments. The participant thanks his triathlon teammates for welcoming, supporting, and cheering him on. The results of the present study do not constitute endorsement by the American College of Sports Medicine.

## Data Availability

The datasets generated and analyzed during the current study are not publicly available to protect the privacy of the participant but are available from the corresponding author upon reasonable request.

Supplemental Digital Content 1.pdf

