## Supplemental Figures 9 and 10 for "One year of testosterone therapy in a transmasculine amateur triathlete: changes in hormone cycles and exercise capacity"

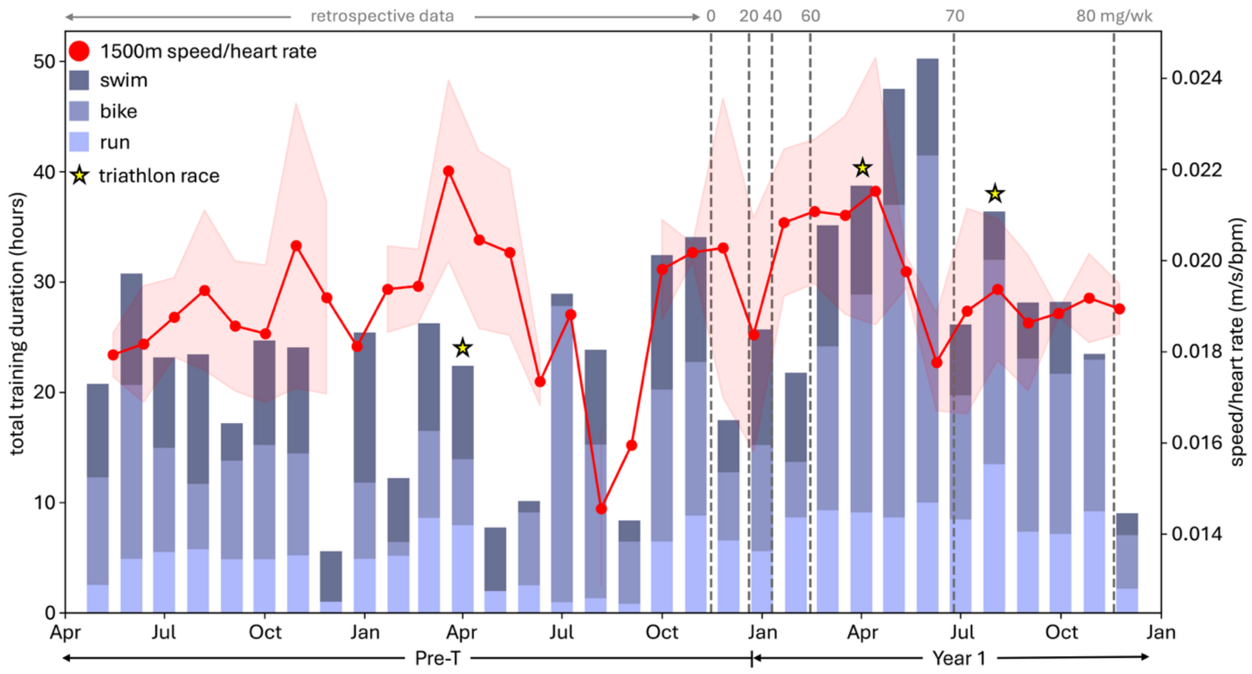


**Figure 9.** Triathlon training volume and fitness calculated from smartwatch data. Retrospective Garmin smartwatch data was collected from 1.5 years prior to the start of the study. On the left axis, total hours of swimming, outdoor biking, and running are plotted in dark blue, medium blue, and light blue, respectively. Weekly stationary bike rides ranging from 45-75 minutes were not recorded. On the right axis, the mean and standard deviation of the participant's fastest 1500m split each week divided by his average heart rate for that run are calculated across four week intervals and plotted in red. From May-September Pre-T, outside factors limited the participant's ability to train. Both speed/heart rate peaks, March Pre-T and March Year 1, correspond to the participant peaking for the April races. Total training hours increased prior to the August race in Year 1. Testosterone dosage level changes are marked by a dashed gray line with the total injection amount listed above in mg/wk.

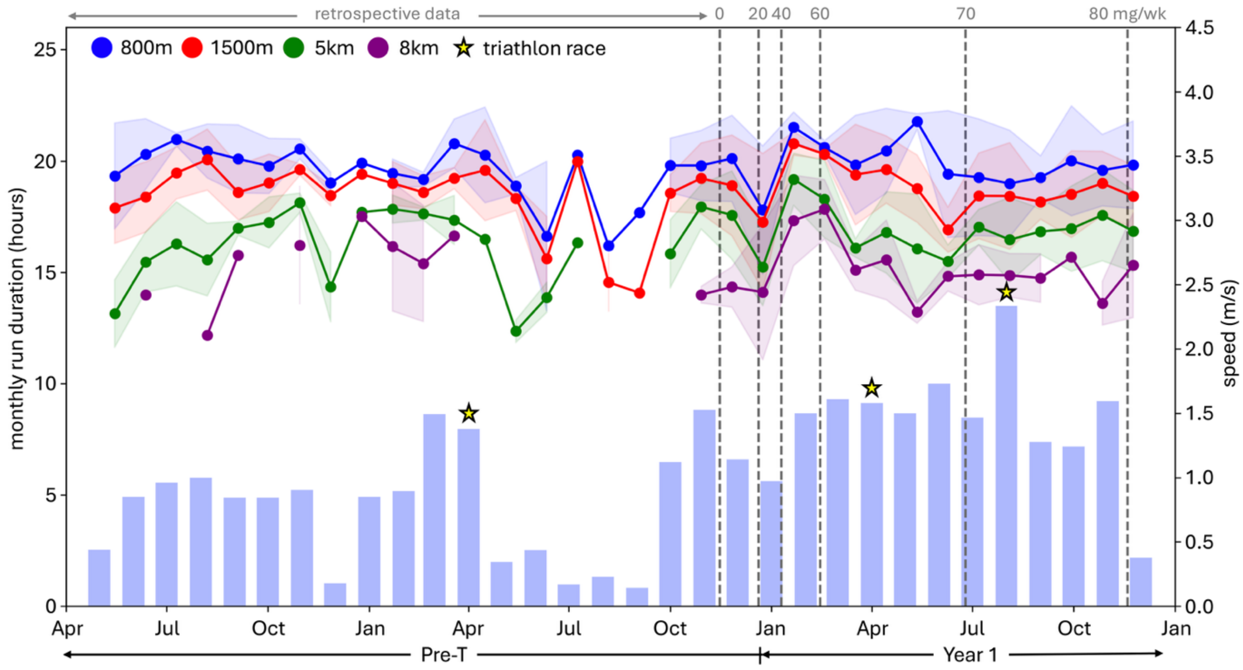


**Figure 10.** The fastest 800m, 1500m, 5km, and 8km run splits each week extracted from the smartwatch data. The mean, shown with dots, and standard deviation, shown with shaded areas, was calculated across four week intervals. The speed of each of these distances is plotted on the right axis in red, blue, green and purple, respectively. On the left axis, total monthly run duration is plotted in hours. All data was extracted from the participant's smartwatch. Testosterone dosage level changes are marked by a dashed gray line with the total injection amount listed above in mg/wk.
